# Economic evaluation of non-pharmacological interventions for fatigue in patients with long-term medical conditions

**DOI:** 10.1101/2025.08.01.25332798

**Authors:** Mon Mon Yee, Christopher Burton, Joanna Leaviss, Jessica E. Forsyth, George Daly, Sarah Davis

**Author notes:** **Corresponding Author:** Mon Mon Yee.

## Abstract

**Background:** Persistent fatigue is a frequent symptom in chronic medical conditions. Systematic reviews of non-pharmacological interventions for fatigue have identified interventions that are effective at reducing fatigue, but there is limited published evidence on the cost-effectiveness of these interventions.

**Objective:** To identify non-pharmacological fatigue interventions that have the potential to be cost-effective and would warrant further investigation in future cost-effectiveness studies.

**Design:** Decision-analytic modelling with quality-of-life outcomes mapped from a systematic review and network meta-analysis of fatigue outcomes and intervention costs estimated from staff time.

**Setting:** UK National Health Service (NHS)

**Participants:** People with persistent fatigue associated with a chronic medical condition

**Interventions:** Non-pharmacological fatigue interventions versus usual care

**Primary and secondary outcome measures:** Net monetary benefit from a UK NHS and Personal Social Services (PSS) perspective; quality-adjusted life years (QALYs) gained; intervention costs valued at 2022/23 prices; costs and benefits discounted at 3.5% per annum.

**Results:** In the base case analysis, expected costs from the probabilistic analysis for individual and group interventions were: £267 and £157 for physical activity promotion, £810 and £485 for CBT-Fatigue, and £462 and £214 for mindfulness. The expected QALYs gained were similar for mindfulness and physical activity promotion (0.061 and 0.060 respectively), but lower for CBT-Fatigue (0.045). All interventions provided positive incremental net monetary benefit (INMB) versus usual care when valuing a QALY at £20,000. However, since group interventions are less costly than individual ones, and we assumed equivalent clinical benefit, they are expected to provide greater INMB. These findings remained robust across different scenarios, except for CBT-Fatigue (individual) which had negative INMB in some scenarios.

**Conclusions:** CBT-Fatigue, physical activity promotion and mindfulness interventions all demonstrated the potential to be cost-effective versus usual care. Future research is recommended to compare the cost-effectiveness of these interventions across a broad population with different chronic conditions.

**STRENGTHS AND LIMITATIONS OF THIS STUDY:**

- Fatigue outcomes were estimated from a robust systematic review and network analysis that pooled data across studies conducted across multiple chronic conditions, with multiple sclerosis being most common.
- The treatment effects for interventions delivered to groups and individuals are assumed to be similar, but this assumption was only supported by an analysis exploring separate treatment effects for group and individual CBT-Fatigue interventions.
- The results for group physical activity promotion and individual mindfulness interventions should be treated with caution owing to there only being data beyond end of treatment for individual physical activity promotion and group mindfulness interventions.
- Healthcare costs were restricted to staffing costs for delivering the intervention and therefore do not capture any impact on resource use outside of the intervention or any non-staff intervention cost such as access to a specific digital tool.

## INTRODUCTION

Persistent fatigue is common in long-term medical conditions.^1^ Chronically-ill patients usually describe fatigue as “more than ordinary tiredness” that could have an impact on their quality of life.^2^ Fatigue is often overlooked and can persist even after the underlying disease has been fully controlled.^3^ The experience of fatigue appears to show similarities across different conditions.^3^ Currently, there are no licensed pharmacological treatments for fatigue in chronic conditions. Several non-pharmacological interventions have been developed in order to address fatigue in such conditions, targeting physical or psychological aspects, or a combination of both.

This research has two main aims: to explore the potential cost-effectiveness of non-pharmacological fatigue interventions and to identify the interventions that would warrant further investigation in future cost-effectiveness studies.

## METHODS

This work was conducted as a part of an evidence synthesis examining both the clinical and cost-effectiveness of non-pharmacological interventions for fatigue in long-term medical conditions.^4^ The review of clinical effectiveness studies including the network meta-analysis (NMA) and the review of cost-effectiveness studies are reported elsewhere.^5^ The NMA of randomised controlled trials (RCTs) examined outcomes at three time points: end of treatment (EOT), short-term (ST) which was defined as up to 3 months after EOT, and long-term (LT) defined as more than 3 months after EOT. There was a large diversity of non-pharmacological interventions reported across the RCTs included in the clinical effectiveness review. These were grouped into intervention categories to allow evidence to be pooled across studies which were considered to have interventions that were sufficiently similar. A large number of interventions were identified and included in the NMA, and many of these failed to demonstrate treatment effects that persisted beyond EOT. We therefore chose to focus the *de novo* economic analysis on those interventions which had the strongest evidence for clinical effectiveness. Consequently, we selected interventions for inclusion in the *de novo* economic evaluation which reported fatigue outcomes at LT follow-up and which demonstrated a statistically significant difference in treatment effect compared to usual care in the NMA for either the ST or LT follow-up. The expected health outcomes and expected costs were estimated relative to usual care in order to align with the approach taken in clinical effectiveness review and NMA. The cost-effectiveness analyses were conducted from the perspective of UK National Health Service (NHS) and personal social services (PSS). Costs are reported in UK pound sterling, valued at 2022/23 prices and costs and benefits were discounted at 3.5% per annum. The data sources and assumptions included in the economic model were informed by discussion with clinical experts and patient and public involvement experts who were able to draw on both their own lived experience and findings from focus groups conducted with people with fatigue associated with long-term conditions.

### Health outcome measures

Only a minority of studies included in the NMA reported a measure of health utility directly. Therefore, we applied a mapping algorithm to estimate utility values from the fatigue outcomes estimated by the evidence synthesis for each intervention category over which effectiveness estimates had been pooled.

#### Identification and selection of mapping algorithms

A targeted literature review was conducted to identify papers which describe statistical mapping methods to convert fatigue-specific patient reported outcome measures used in chronic health conditions to health state utility values derived from generic preference-based measures (PBM), such as the EQ-5D and SF-6D. The detailed methods are provided in the supplementary appendix.

Our electronic database searches identified 96 papers. A study by Goodwin *et al*.^6^ that mapped from the Fatigue Severity Scale (FSS) to health state utility values using EQ-5D-3L, SF-6D and MSIS-8D was identified from both electronic database and HERC database searches. After removing duplicates and reviewing the papers, two additional studies^7-8^ were found mapping CIS-F to EQ-5D-5L and fatigue measured on a visual analogue scale (VAS) to EQ-5D-3L, respectively. These three papers are summarised in the supplementary appendix (Table S1). No further relevant studies were identified from our citation search. Of these three papers, Goodwin *et al.* was selected in preference to the remaining papers because it was the only included study that mapped from FSS, which is frequently used to measure fatigue severity, to PBMs. The studies by Eriksson *et al*.^7^ and Bloem *et al*.^8^ were considered less appropriate than the study by Goodwin *et al*. because they included other patient characteristics (e.g. disease severity measures, comorbidities, depression and anxiety scores) in their regressions which may not be available in the studies included in our evidence synthesis. Goodwin *et al.* used five regression models to map from either FSS total score or FSS item score to each PBM (EQ-5D-3L, SF-6D and MSIS-8D) using both ordinary least squares (OLS) and censored least adjusted deviation (CLAD) specifications. The regression model mapping from total FSS to SF-6D was reported as performing better than the regression model mapping to EQ-5D. The MSIS-8D mapping algorithm was considered less useful for estimating utilities in populations with chronic conditions other than MS. Although the algorithm mapping to SF-6D was derived in an MS population, we felt this was broadly applicable to other populations of patients with chronic conditions as both the FSS and the SF-6D are generic tools that have been validated for use across a range of health care conditions.^9^ After considering the mapping studies identified in this review, we chose to use the regression model mapping from FSS to SF-6D provided by Goodwin *et al.* in our *de novo* economic model.

#### Quality-adjusted life-years

The QALY gains for each intervention and usual care were estimated using an area under the curve approach based on the average time points for fatigue outcomes reported in the studies contributing to the NMA. As the absolute utility values were not reported consistently across studies, we have used data on the difference in fatigue scores between arms, which were then converted to absolute FSS scores and mapped to SF-6D values to estimate the QALYs gained relative to the usual care.

For each intervention category, the studies contributing to that intervention category in the NMA were used to estimate the timing for EOT and the two follow-up points (ST and LT) used when estimating QALYs. For studies that have reported fatigue scores for two different LT follow-ups, the base-case NMA included the longest follow-up data but a sensitivity analysis was conducted incorporating the earliest LT follow-up point (i.e. closest to 3 months after EOT). This was done to ensure that any waning of treatment effect over time did not bias the analysis against studies that had a greater duration of follow-up. We explored the impact of using the data from this NMA sensitivity analysis in the economic analysis (scenario analysis) and adjusted the timing of the LT follow-up accordingly. The details of this NMA scenario analysis can be found in Table S9 of the supplementary appendix.

It was assumed that the baseline FSS score would be the same across interventions and the usual care comparator arm and this was based on an estimate from Goodwin *et al.^6^* (43.73, 95%CI 14.13 to 73.33). The absolute FSS score for usual care was kept fixed at its baseline value for all follow-up time points. For the intervention arms, the standardised mean differences (SMDs) of fatigue scores provided by the NMA across all conditions for each intervention relative to the usual care were converted to a mean difference on the FSS scale using the standard deviation (SD) of baseline FSS score of 15.1 reported by Goodwin *et al.* in the base case. The SD of FSS scores derived from studies included within the NMA was used in the scenario analysis (see supplementary appendix for further details). This difference in fatigue scores between intervention and usual care was used to estimate the absolute FSS score after baseline for each intervention at each follow-up point (absolute FSS score = difference in FSS score + baseline FSS score). It did not seem reasonable to assume a sudden return to baseline fatigue scores after the last follow-up. Therefore, it was assumed that interventions would experience a linear decline in treatment effect between the last follow-up and 24 months after baseline in the base case. This assumption was informed by LT follow-up data from two studies which showed the potential for treatment effects to be maintained beyond one year.^10-11^ No further treatment effect was assumed to persist beyond that point. If data were missing at the ST follow-up point, a linear change in FSS between the EOT and LT follow-up points was assumed. This assumption was tested in the scenario analysis, where the treatment effect was assumed to be zero when data was unavailable. QALYs were discounted using a discount rate of 3.5% per annum, as recommended by NICE.^12^ The time horizon was 24 months for the base-case analysis and 15 and 48 months in the scenarios exploring pessimistic and optimistic assumptions, respectively, regarding the persistence of treatment effects.

### Resource use and costs

The studies included in the NMA were examined to determine the resources required to deliver the interventions included within each intervention category for the evidence synthesis. Expected costs were estimated separately for interventions delivered to groups versus those delivered to individuals as the latter are usually cheaper on a cost per patient basis. Information was extracted on the number of sessions, duration of sessions and the health-care professionals involved in delivering or facilitating the interventions. For interventions delivered to groups, information was also extracted on the number of individuals who started the intervention, the group size and the number of groups (assumed to be one if not stated) to allow an estimation of the average cost per patient. Unit costs were taken from the Personal Social Services Research Unit (PSSRU) Unit Costs of Health and Social Care 2023^13^ with the exception of cognitive behavioural therapy (CBT), which was only reported in a previous edition of the PSSRU Unit Costs (2017),^14^ and therefore, this cost was uplifted to 2023 prices using the Hospital and Community Health Service Pay and Prices index based on PSSRU 2023. Unit costs used to estimate the expected intervention costs are summarised in the supplementary appendix (Table S2). Detailed assumptions used in the costing analysis on a study-by-study basis are provided in Tables S3-S8 of the supplementary file. Median costs across each intervention category were used in the economic analysis for the deterministic analysis. Interventions were typically shorter than 1 year so discounting was not applied.

The costing analysis includes only intervention costs and does not estimate its impact on other healthcare resource use such as reduced primary or secondary care attendances resulting from patients experiencing lower fatigue or increased use from potential adverse effects. Studies identified in the cost-effectiveness review which assessed the overall impact of an intervention on healthcare costs have been reported elsewhere.^5^

### Cost-effectiveness

The cost-effectiveness analysis was not taken on a study-by-study basis. Instead we focused on intervention categories that have been considered sufficiently similar to be analysed together within the NMA. The same effectiveness was applied across group and individual interventions in the base case as these were pooled within the NMA. A scenario analysis was conducted to test the validity of assuming equivalent treatment effects for group and individual interventions, using data for CBT-Fatigue, where sufficient data were reported to estimate treatment effects separately (see Table S10 in the supplementary appendix for detailed NMA results). The net monetary benefit (NMB) was estimated based on the willingness to pay threshold of £20,000 per QALY gained. The cost-effectiveness estimates were generated using both deterministic and probabilistic versions of the model. The deterministic model applied the point estimates of the parameters, whereas the probabilistic model used Monte Carlo sampling across 5,000 iterations to generate distributions of expected health outcomes and costs for each treatment group. Uncertainty around treatment effect was handled using Convergence Diagnostics and Analysis (CODA) samples while baseline FSS score and regression coefficients were sampled from a normal distribution and the intervention costs from gamma distributions. When the variance-covariance matrices for the mapping coefficients were not reported in the paper, it was assumed that the coefficient for FSS was independent of the intercept parameter. As standard errors (SEs) of costs were unavailable, they were estimated using the lowest and highest expected costs of each intervention type across relevant studies. When the intervention cost was limited to a single study, the SE was assumed to be equal to 25% of the expected cost.

Scenario analyses were also conducted exploring the impact of (i) optimistic and pessimistic durations for treatment effect decline, (ii) alternative baseline FSS score and SD for FSS based on studies from the NMA EOT network, (iii) different SMDs for FSS between individual and group CBT-Fatigue interventions, (iv) alternative SMDs using the shorter follow-up point in the LT network, (v) assuming that SMD of mindfulness at the ST follow-up timepoint is zero by removing the linear change assumption between EOT and LT, and (vi) lower and upper costs.

## RESULTS

Based on the NMA results, physical activity promotion, CBT-based fatigue interventions (CBT-Fatigue) and mindfulness were found to be eligible for our economic analysis as they had RCTs reporting fatigue outcome at more than 3 months after the EOT and they had a statistically significant SMD in fatigue scores versus usual care at either ST or LT follow-up. We chose not to include remote ischaemic conditioning in the economic analysis, as although this did achieve a statistically significant difference in fatigue versus usual care in the LT NMA, this was based on a single study in stroke patients and the intervention was potentially specific to the population and so less likely to be generalisable to patients experiencing fatigue associated with other chronic conditions.

### Health outcomes

#### Quality-adjusted life-years

The mapped SF-6D utility values assumed in the base-case analysis for interventions and the usual care are shown in Figure 1 and those used in the scenario analyses are presented in Figure S1-S4 of the supplementary file. Physical activity promotion was estimated to have the highest utility values at ST and LT follow-up timepoints in both base case and sensitivity analyses. Both probabilistic and deterministic base cases indicated that mindfulness and physical activity promotion interventions were associated with higher QALYs gained compared to CBT-Fatigue, with mindfulness slightly exceeding physical activity promotion (see Table 1).

**Figure 1.**
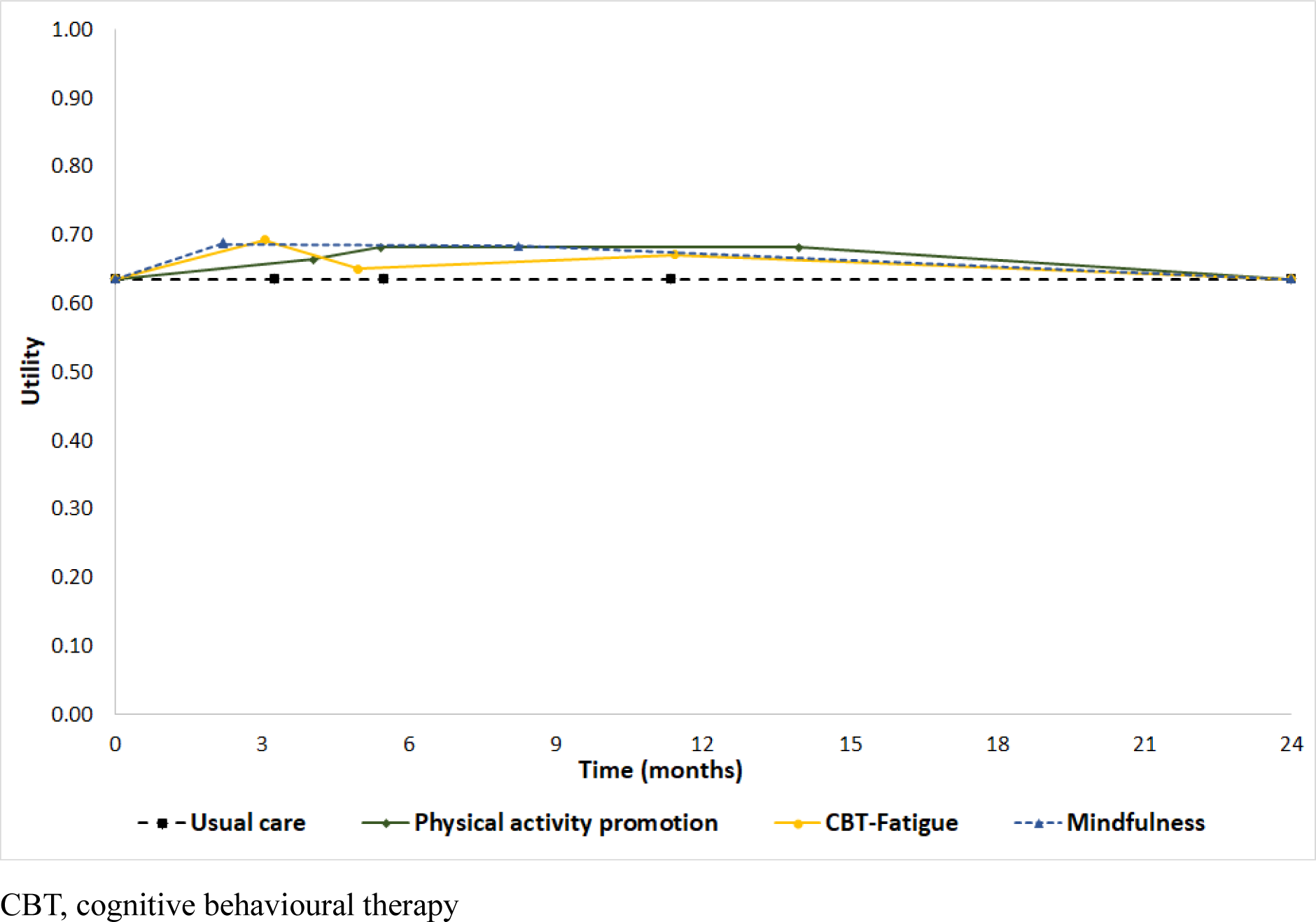
Utility values of interventions across different timepoints.

**Table 1.** Base-case cost-effectiveness results.

| Intervention type | Group or individual intervention | Number of study arms | Median cost (£) | Mean QALYs (discounted) | Incremental Costs | Incremental QALYs | NMB at £20,000 threshold | INMB against UC |
| --- | --- | --- | --- | --- | --- | --- | --- | --- |
| **Base-case (deterministic)** |  |  |  |  |  |  |  |  |
| Usual care | - | - | £0 | 1.227 | - | - | £24,531 | - |
| Physical activity promotion | Individual | 3 | £267 | 1.287 | £267 | 0.060 | £25,465 | £934 |
| CBT-Fatigue | Individual | 10 | £817 | 1.271 | £817 | 0.045 | £24,606 | £75 |
| Mindfulness | Individual | 2 | £465 | 1.287 | £465 | 0.060 | £25,272 | £741 |
| Physical activity promotion | Group | 2 | £156 | 1.287 | £156 | 0.060 | £25,576 | £1,045 |
| CBT-Fatigue | Group | 5 | £484 | 1.271 | £484 | 0.045 | £24,939 | £408 |
| Mindfulness | Group | 1 | £215 | 1.287 | £215 | 0.060 | £25,522 | £991 |
| **Base-case (probabilistic)** |  |  |  |  |  |  |  |  |
| Usual care | - | - | £0 | 1.223 | - | - | £24,463 | - |
| Physical activity promotion | Individual | 3 | £267 | 1.283 | £267 | 0.060 | £25,397 | £934 |
| CBT-Fatigue | Individual | 10 | £810 | 1.268 | £810 | 0.045 | £24,553 | £90 |
| Mindfulness | Individual | 2 | £462 | 1.284 | £462 | 0.061 | £25,222 | £760 |
| Physical activity promotion | Group | 2 | £157 | 1.283 | £157 | 0.060 | £25,508 | £1,045 |
| CBT-Fatigue | Group | 5 | £485 | 1.268 | £485 | 0.045 | £24,878 | £415 |
| Mindfulness | Group | 1 | £214 | 1.284 | £214 | 0.061 | £25,470 | £1,007 |

QALY, quality-adjusted life years; NMB, net monetary benefit; INMB, incremental net monetary benefit; UC, usual care; CBT, cognitive behavioural therapy

#### Resource use and costs

A cost was derived from at least one arm of twenty-one studies that included a total of twenty-three intervention arms that were classified as CBT-Fatigue, physical activity promotion or mindfulness. Costs could not be estimated for five studies (Nguyen *et al*.^15^, Kucharski *et al*.,^16^ Okkersen *et al*.^17^, Pottgen *et al*.^18^ and Katz *et al*.^19^) because no information was provided in the publications on the duration of sessions with healthcare providers or the interventions included only web-based tools for which no cost data were available. The range of costs for each intervention is presented in Figure 2 as box-and-whisker plots. As summarised in Table 1, the mean costs estimated by the probabilistic model were similar to the median costs from the deterministic, suggesting that group physical activity promotion incurred the lowest costs, whereas individual CBT-Fatigue had the highest costs due to higher staff contact time in terms of frequency and duration of sessions.

**Figure 2.**
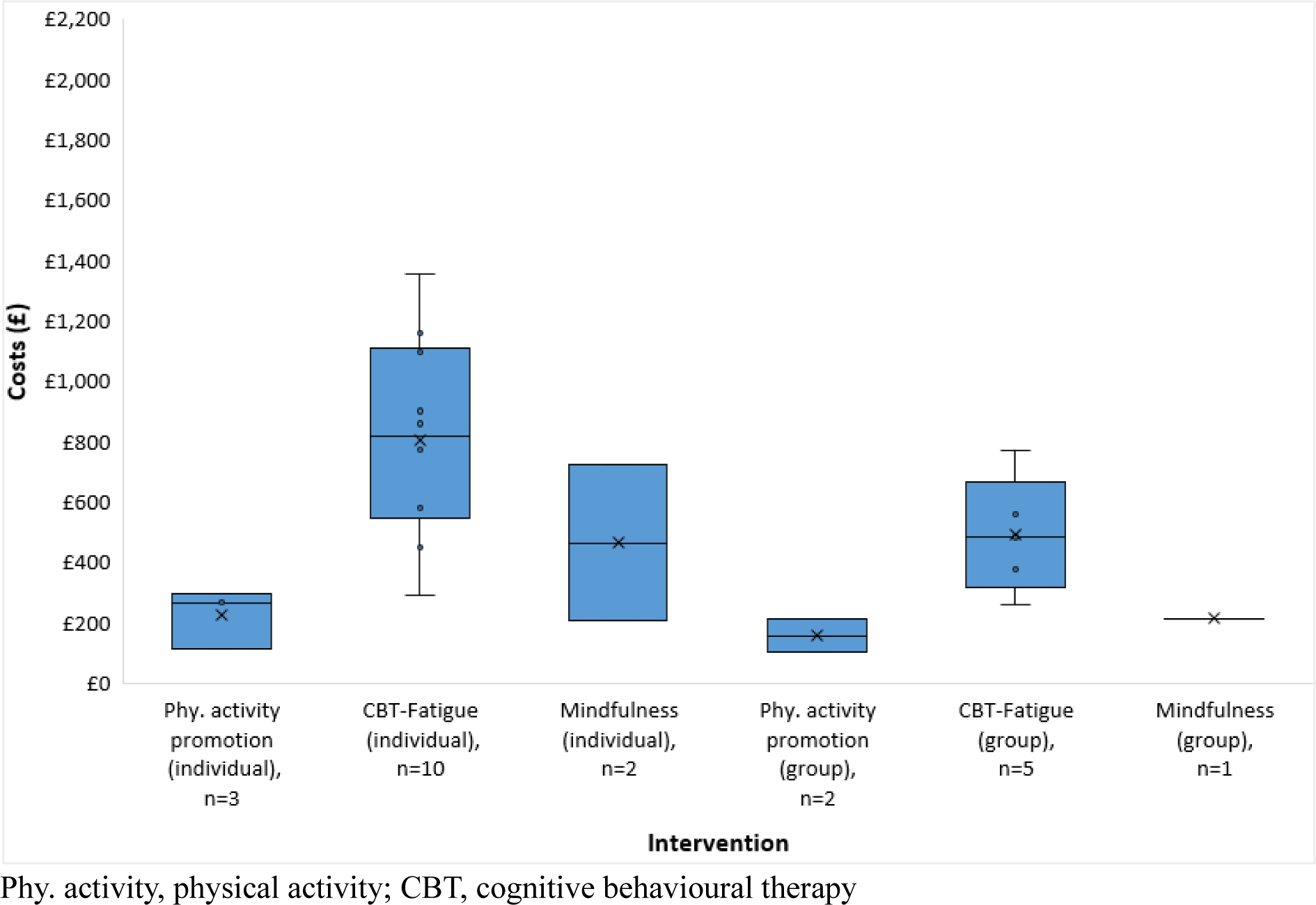
Distribution of costs by type of intervention.

#### Cost-effectiveness

The incremental costs and QALYs and the incremental net monetary benefit (INMB) for each intervention versus usual care, using deterministic and probabilistic models, are presented in Table 1. It can be seen that for all interventions, the INMB versus usual care is positive, which means that the monetary value of the QALYs gained is greater than the intervention cost, indicating that the intervention would be considered cost-effective when valuing a QALY at £20,000. In both deterministic and probabilistic analyses, physical activity promotion (group intervention) is estimated to provide the highest INMB compared to usual care, followed by group mindfulness, and then individual physical activity promotion. The higher cost of CBT-Fatigue interventions mean that they have the lowest INMB versus usual care with individual CBT-Fatigue having the lowest INMB. The results of scenario analyses (see Table S11 of the supplementary file) are similar to the base case for the majority of the interventions, with the exception being CBT-Fatigue (individual intervention). Due to the higher cost of this intervention, it has a negative INMB, indicating an incremental cost-effectiveness ratio (ICER) greater than £20,000 per QALY versus usual care under certain conditions: (i) pessimistic assumption of the treatment effect decline, (ii) alternative baseline FSS score and (iii) higher intervention costs.

## DISCUSSION

There have been a number of trial-based economic evaluations of non-pharmacological interventions for fatigue in specific medical conditions.^11,20-22^ To our knowledge, no study has assessed the cost-effectiveness of such interventions across multiple conditions using decision analytic modelling. Our *de novo* economic assessment evaluated the cost-effectiveness of interventions that had been found, in a systematic review and NMA of interventions across long-term medical conditions, to demonstrate a statistically significant improvement in fatigue beyond the end of treatment. The base-case model suggests that physical activity promotion, CBT-Fatigue and mindfulness could all be cost-effective at the willingness-to-pay threshold of £20,000 per QALY gained, regardless of whether they are delivered individually or in groups. These findings were robust under the scenario analyses conducted with the exception of CBT-Fatigue delivered to individuals, which had the lowest INMB in the base-case and was therefore less robust to scenarios with more pessimistic assumptions.

### Strengths and limitations

One of the key strengths of our analysis is that it was based on the pooled results of fatigue outcomes across multiple long-term conditions, derived from a comprehensive systematic review and NMA. Studies on multiple sclerosis accounted for around half of the included studies in each network. Both base case and scenario analyses are probabilistic, capturing the model parameter uncertainty. In addition, the probabilistic model explicitly incorporated the correlation between efficacy estimates (fatigue outcomes) using the CODA samples generated by the NMA.

Our economic assessment is also subject to some limitations. Very few studies reported the UK estimates of resource use other than those associated with delivering the intervention and therefore, the estimated incremental costs might be overestimated or underestimated if the interventions result in decrease or increase in other resource use. Additionally, a detailed cost breakdown for internet or app based tools (e.g., MS Invigor8^20^, ELEVIDA^18^ Tailorbuilder^23^) were unavailable; therefore, the cost of such tools were excluded from the model, which might lead to an underestimation of costs for some interventions. In terms of intervention costs per patient, interventions delivered to groups were found to be less costly than the same type of interventions delivered individually. For group interventions, group size determined the cost per patient. Where the group size was not reported in the studies, it was assumed that there was a single group and that may have underestimated the cost per patient if smaller groups were used instead.

There is a broad range of costs for each type of intervention across included studies, indicating high level of heterogeneity in expected cost estimates. Sensitivity analyses explored the impact of the heterogeneity in intervention costs, and only CBT-Fatigue (individual) was found to have an ICER greater than £20,000 per QALY versus usual care when the highest cost was applied. This finding aligns with the results from the within-trial economic evaluations reported by Thomas *et al.,^21^* Chong *et al.,^22^*and Hewlett *et al*.^11^ where CBT was dominated by usual care because of higher intervention costs and small negative incremental QALYs. Chong *et al*.^22^ compared physical activity promotion to CBT directly using a within-trial analysis of the RCT (reported by Bachmair *et al*.^24^) and their conclusion that CBT was dominated by physical activity promotion is replicated in our *de novo* analysis informed by effectiveness data from multiple studies. Given that CBT-Fatigue may include elements of physical activity promotion or mindfulness, these findings raise the question of whether there is “added value” from the additional cognitive components of CBT-Fatigue interventions, given that these are generally more intensive to deliver.

The expected QALYs were based on the assumption that the difference between interventions and usual care would start to decline after the last follow-up timepoint and become zero at 24 months (from the baseline). This assumption was tested using optimistic and pessimistic scenarios regarding the treatment effect dissipation. All interventions were less cost-effective when making a more pessimistic assumption about the persistence of the treatment effect but only CBT-Fatigue (individual) had an ICER greater than £20,000 per QALY when assuming that the treatment effect declined rapidly within a year after the intervention. Another limitation is that the probabilistic model had to use SEs for mapping coefficients when sampling from a normal distribution instead of using variance-covariance matrix, which might overestimate the uncertainty of the mapping algorithm.

The scenario analysis exploring the impact of allowing for different treatment effects for CBT-Fatigue from group and individual interventions had minimal impact on the cost-effectiveness estimate for CBT-Fatigue versus usual care due to prediction of similar treatment effects. However, it should be noted that we were unable to conduct a similar analysis exploring the impact of allowing for different treatment effects of group and individual interventions for the mindfulness and physical activity promotion. This was due to there being only being data beyond the end of treatment available for group mindfulness interventions and individual physical activity promotion interventions. As such the cost-effectiveness results for individual mindfulness interventions and group physical activity promotion interventions should be interpreted with caution.

Although our analysis is able to estimate which interventions have the highest INMB, any head-to-head comparison should be interpreted with caution given the heterogeneity in intervention costs across the included studies as shown by the overlapping confidence intervals for the intervention costs (see figure 2). However, across all the scenario analyses explored, CBT-Fatigue had lower QALY gains than mindfulness or physical activity promotion due to a smaller and non-statistically significant difference in fatigue scores at the ST follow-up. Although it should also be noted that no efficacy data were available for mindfulness from the ST follow-up and the assumption of a linear change in fatigue between the EOT and the LT follow-up point for the mindfulness intervention could be optimistic. However, in our scenario analysis we found that mindfulness had greater QALY gains than CBT-Fatigue even when assuming no clinical efficacy at the short-term follow-up point for mindfulness. The analysis for CBT-Fatigue was also the only analysis in which there was data from multiple studies at both the ST and LT follow-up points and in which the studies informing these endpoints were conducted across more than one type of chronic condition (nine studies for multiple sclerosis, three for stroke, three for musculoskeletal disease and one for inflammatory bowel disease). Therefore, the greater QALY gains for mindfulness and physical activity promotion in this analysis should not be overinterpreted as demonstrating clinical superiority for patients with chronic conditions, as the LT data for these interventions are based on a smaller set of studies covering a limited set of conditions (2 studies for physical activity promotion in musculoskeletal conditions and 1 study for mindfulness in multiple sclerosis for mindfulness).

## CONCLUSIONS

This *de novo* economic evaluation indicates that mindfulness interventions, physical activity promotion interventions and CBT-fatigue interventions have the potential to be a cost-effective means for improving quality of life in people experiencing fatigue associated with a chronic condition when compared to usual care. If it is assumed that group and individual interventions have similar efficacy, as supported by our analysis of CBT-Fatigue interventions, then group interventions tend to be lower cost to deliver and are therefore more cost-effective than individual interventions. Whilst CBT-Fatigue had higher costs and lower QALY gains than the other interventions, the clinical effectiveness estimates for the other interventions are based on fewer studies conducted across a narrower range of conditions. We therefore recommend that future research is conducted to compare the cost-effectiveness of CBT-Fatigue, physical activity promotion and mindfulness interventions across a broad population with different chronic conditions.

## Acknowledgements

The authors would like to thank Helen Dawes, Vincent Deary, Julia Newton, Kate Fryer, Samantha McCormick and David Coyle for their advice on the data sources and assumptions informing the economic analysis and the interpretation of the results. We would like to thank Kate Ren, Professor of Statistical Health Technology Assessment at the University of Sheffield, who provided statistical advice and guidance to JEF and GD when conducting the network meta-analysis and analysis of baseline fatigue scores.

## Author Contributions

MMY and SD conceptualised and designed the economic evaluation. JL, JEF, GD and CB conducted the systematic review and network meta-analysis that informed the economic analysis and provided advice on the incorporation of clinical efficacy evidence within the analysis. All authors contributed to development, analysis, writing and editing the manuscript. All authors read and approved the final version.

## Patient and public involvement

The research group included two members with relevant lived experience and an academic expert in patient and public involvement who were involved in discussion regarding the data sources and assumptions for the economic modelling. Their contributions were informed by 5 focus groups convened of people with fatigue associated with long term conditions.

## Patient consent for publication

Not applicable.

## Ethics approval

Not applicable.

## Provenance and peer review

Not commissioned; externally peer reviewed.

## Data availability statement

All data relevant to the study are included in the article or uploaded as supplementary file. The economic model is available from the corresponding author upon reasonable request

## Supplementary material

This content has been supplied by the author(s).

