## Supplementary data for "Economic evaluation of non-pharmacological interventions for fatigue in patients with long-term medical conditions"

### SUPPLEMENTARY FILE

#### Identification and selection of mapping algorithms

In order to be eligible for our review, mapping studies had to be conducted in a population of adults who have a long-term medical condition which is stable regardless of treatment. Studies that included cancer, medical conditions reliant on fatigue for diagnosis (e.g., fibromyalgia, myalgic encephalomyelitis or chronic fatigue syndrome), medical conditions arising from infection (e.g., long COVID or post-viral illness) or injuries were excluded from the review as in the clinical effectiveness study. Studies that report mapping algorithms from disease-specific instruments, where fatigue was reported only as one of the domains, were not considered to be useful as data on the other domains would be unlikely to be available across the broad range of studies included in the NMA. The searches were performed in four electronic databases (MEDLINE, PubMed, Web of Science and the Cumulative Index to Nursing and Allied Health Literature [CINAHL]) between April 2024 and June 2024. The search strategies (described below) were based on a published systematic review of mapping studies and these were adapted to include a fatigue term. We also searched the Health Economic Research Center (HERC) database (version 9.0, last updated on the 1st of December 2023)[1] which is a database of mapping algorithms for calculating EQ-5D from other generic or disease-specific HRQoL instruments. The database is informed by a systematic review of mapping studies which is regularly updated.[2] In addition, we hand-searched the citations and reference lists of relevant papers in PubMed and Google Scholar to identify any further papers which discussed mapping algorithms to convert fatigue-specific measurements to EQ-5D or SF-6D.

**Electronic database search strategies for mapping studies**

*Ovid MEDLINE(R) Epub Ahead of Print and In-Process, In-Data-Review & Other Non-Indexed Citations and Daily*

22nd April 2024

| # | Searches | Results |
|---|---|---|
| 1 | preference based.tw. | 1814 |
| 2 | (multiattribut or multi attribute or mau or maui).tw. | 2460 |
| 3 | (euroqol or euro qol or eq5d or eq 5d).tw. | 17955 |
| 4 | (sf 6d or sf6d or short form 6d or shortform 6d or sf six or sfsix or shortform six or short form six).tw. | 1079 |
| 5 | health utilities index.tw. | 814 |
| 6 | (quality adj2 (wellbeing or well being)).tw. | 3120 |
| 7 | qwb.tw. | 219 |
| 8 | (assessment adj2 quality adj2 life instrument).tw. | 25 |
| 9 | "aqol*".tw. | 320 |
| 10 | 1 or 2 or 3 or 4 or 5 or 6 or 7 or 8 or 9 | 25717 |
| 11 | (mapping* or mapped or map).tw. | 443444 |
| 12 | (crosswalk* or cross walk*).tw. | 842 |

| | | |
|---|---|---|
| 13 | (estimat* or predicted or predicting or prediction or transfer or transformation or derive or derived or deriving or derivation).ti. | 582141 |
| 14 | 11 or 12 or 13 | 1014667 |
| 15 | "fatigue*".ti. | 34927 |
| 16 | 10 and 14 and 15 | 5 |

*Web of Science: Clarivate Analytics*
7th June 2024

| # | Searches | Results |
|---|---|---|
| #1 | TS=("preference based") | 5850 |
| #2 | TS=(multiattribut or "multi attribute" or mau or maui) | 22298 |
| #3 | TS=(euroqol or "euro qol" or eq5d or eq 5d) | 26492 |
| #4 | TS=("sf 6d" or sf6d or "short form 6d" or "shortform 6d" or "sf six" or sfsix or "shortform six" or "short form six") | 1659 |
| #5 | TS=("health utilities index") | 1133 |
| #6 | TS= ((quality NEAR/2 wellbeing)) OR TS= ((quality NEAR/2 well being)) | 8715 |
| #7 | TS= (qwb) | 304 |
| #8 | TS= ((assessment NEAR/2 quality NEAR/2 life instrument)) | 2857 |
| #9 | TS= (aqol*) | 494 |
| #10 | TS=((15d or "15dimensional" or "15 dimensional")) | 9018 |
| #11 | #1 OR #2 OR #3 OR #4 OR #5 OR #6 OR #7 OR #8 OR #9 OR #10 | 74943 |
| #12 | TS=((mapping* or mapped or map)) | 3635581 |
| #13 | TS=((crosswalk* or cross walk*)) | 59397 |
| #14 | TI=(estimat* or predicted or predicting or prediction or transfer or transformation or derive or derived or deriving or derivation ) | 4509013 |
| #15 | #12 OR #13 OR #14 | 8050668 |
| #16 | TS=(fatigue*) | 996729 |
| #17 | #11 AND #15 AND #16 | 83 |

*Cumulative Index to Nursing and Allied Health Literature (CINAHL): EBSCO*
7th June 2024

| # | Searches | Results |
|---|---|---|
| S1 | TI preference based OR AB preference based | 3,572 |
| S2 | TI ( (multiattribut or multi attribute or mau or maui) ) OR AB ( (multiattribut or multi attribute or mau or maui) ) | 668 |
| S3 | TI ( euroqol or euro qol or eq5d or eq 5d ) OR AB ( euroqol or euro qol or eq5d or eq 5d ) | 2,840 |
| S4 | TI ( sf 6d or sf6d or short form 6d or shortform 6d or sf six or sfsix or shortform six or short form six ) OR AB ( sf 6d or sf6d or short form 6d or shortform 6d or sf six or sfsix or shortform six or short form six ) | 558 |
| S5 | TI health utilities index OR AB health utilities index | 539 |
| S6 | TI ( quality N2 (wellbeing or well being) ) OR AB ( quality N2 (wellbeing or well being) ) | 2,426 |
| S7 | TI qwb OR AB qwb | 96 |
| S8 | TI assessment N2 quality N2 life instrument OR AB assessment N2 quality N2 life instrument | 141 |
| S9 | TI aqol OR AB aqol | 90 |
| S10 | S1 OR S2 OR S3 OR S4 OR S5 OR S6 OR S7 OR S8 OR S9 | 10,359 |
| S11 | TI ( mapping* or mapped or map ) OR AB ( mapping* or mapped or map ) | 57,752 |
| S12 | TI ( crosswalk* or cross walk* ) OR AB ( crosswalk* or cross walk* ) | 790 |

| | | |
|---|---|---|
| S13 | TI (estimat* or predicted or predicting or prediction or transfer or transformation or derive or derived or deriving or derivation) | 121,021 |
| S14 | S11 OR S12 OR S13 | 177,984 |
| S15 | TI fatigue* | 17,031 |
| S16 | S10 AND S14 AND S15 | 2 |

*PubMed*
7th June 2024

| # | **Searches** | **Results** |
|---|---|---|
| #1 | preference based[Title/Abstract] | 1,943 |
| #2 | multiattribut or multi attribute or mau or maui[Title/Abstract] | 15,921 |
| #3 | euroqol or euro qol or eq5d or eq 5d [Title/Abstract] | 19,647 |
| #4 | sf 6d or sf6d or short form 6d or shortform 6d or sf six or sfsix or shortform six or short form six [Title/Abstract] | 7,746 |
| #5 | health utilities index [Title/Abstract] | 831 |
| #6 | quality N2 (wellbeing[Title/Abstract] OR well being[Title/Abstract]) | 263 |
| #7 | qwb[Title/Abstract] | 221 |
| #8 | assessment N2 quality N2 life instrument [Title/Abstract] | 3 |
| #9 | aqol [Title/Abstract] | 339 |
| #10 | #1 OR #2 OR #3 OR #4 OR #5 OR #6 OR #7 OR #8 OR #9 | 44,657 |
| #11 | mapping*[Title/Abstract] OR mapped[Title/Abstract] OR map[Title/Abstract] | 470,822 |
| #12 | crosswalk*[Title/Abstract] OR cross walk*[Title/Abstract] | 922 |
| #13 | estimat*[Title] OR predicted[Title] OR predicting[Title] OR prediction[Title] OR transfer[Title] OR transformation[Title] OR derive[Title] OR derived[Title] OR deriving[Title] OR derivation[Title] | 742,840 |
| #14 | #11 OR #12 OR #13 | 1,199,929 |
| #15 | fatigue*[Title] | 36,518 |
| #16 | #10 AND #14 AND #15 | 6 |

**Table S 1** Summary of studies which mapped fatigue-specific HRQoL measures to generic non-preference-based measures

| Study | Disease category | Quality of life measures for fatigue | | Mapping models investigated | Regression equation |
|---|---|---|---|---|---|
| | | From | To | | |
| Goodwin *et al.* 2019[3] | Multiple sclerosis (MS) | Fatigue Severity Scale (FSS) | EQ-5D-3L*<br>SF-6D<br>MSIS-8D | OLS model†<br>CLAD model‡ | SF-6D estimate = 0.897–0.006*FSS total score<br>MSIS-8D estimate = 1.084–0.008*FSS total score – 0.001*age – 0.024*gender [0 male, 1 female]<br><br>If age and gender are not available:<br>MSIS-8D estimate = 0.985–0.007*FSS total score |
| Eriksson *et al.* 2019[4] | Multiple sclerosis (MS) | VAS (0 to 10 ) | EQ-5D-3L | OLS model | Regression coefficients are provided for eighteen variables included when using the UK valuation set for EQ-5D (age, MS type, employment status, EDSS score as a categorical variable, Fatigue VAS, Cognition VAS, and two for categories of treatments received). The VAS fatigue measure has a coefficient of -0.024 (95%CI -0.026 to -0.023). |
| Bloem *et al.* 2021[5] | Idiopathic Pulmonary Sclerosis (IPF) | Checklist Individual Strength subscale fatigue (CIS-Fatigue) | EQ-5D -5L | Stepwise multiple regression models | EQ-5D-5L VAS = 80.700 – 0.529*CIS-Fatigue – 1.611*HADS-Anxiety – 5.440* Comorbidities + 0.339*TLCO %Pred<br><br>EQ-5D-5L index value = 0.829 – 0.003*FQL-Exhausted – 0.032*mMRC – 0.011*HADS-Anxiety – 0.014*HADS-Depression + 0.003*TLCO%Pred |
| | Sarcoidosis | Fatigue Quality List (FQL) | | | EQ-5D-5L VAS = 98.672 – 0.815*CIS-Fatigue + 0.340*FQL-Pleasant – 1.387*HADS-Depression<br><br>EQ-5D-5L index value = 0.986 – 0.002*FQL-Exhausted – 0.070*mMRC – 0.019*HADS-Anxiety |

* The regression equation for EQ-5D-3L was not reported in the paper because of poor performance of EQ-5D algorithms.

† Best performing model for SF-6D.

‡ Best performing model for EQ-5D and MSIS-8D.

EQ-5D-3L ,EuroQoL 5 Dimensions-3 levels; EQ-5D-5L ,EuroQoL 5 Dimensions 5 levels; SF-6D,short-form 6 Dimensions, MSIS-8D,Multiple Sclerosis Impact Scale 8 Dimensions; EQ-5D-5L VAS ,EuroQoL 5 Dimensions 5 levels Visual Analog Scale ; VAS, Visual Analog Scale; HADS, Hospital Anxiety and Depression Scale ; TLCO%Pred, Transfer Factor of the lung for carbon monoxide percentage predicted ; mMRC, modified Medical Research Council-Dyspnoea; OLS, ordinary least squares; CLAD, Censored Least Adjusted Deviation

### Detailed costing assumptions

**Table S 2** Unit costs applied in the costing of interventions

| Healthcare staff | Unit cost | Ratio of direct to indirect time | Cost per minute* | Source |
|---|---|---|---|---|
| Physiotherapist | 41 per hour | 1:0.37 | £0.94 | PSSRU 2023[6] |
| CBT | 100 per 55-min session | NA | £2.15 | PSSRU 2017,[7] uplifted to 2022/23 prices |
| Mindfulness-based cognitive therapy | 175 per 2-hour session | NA | £1.72 | PSSRU 2017,[7] uplifted to 2022/23 prices |

* The qualification costs and cost per minute of face-to-face time were included, where available. Otherwise, the ratios of direct to indirect time were used to estimate the cost per minute of face-to-face time.

CBT, cognitive behavioural therapy; PSSRU, Personal Social Services Research Unit

**Table S 3** Costs of physical activity promotion delivered to individuals

| First author and year of publication | Session facilitator | Face to face (F) or distance (D) | Number of sessions | Duration of sessions (minutes) | Unit cost | Cost per patient |
|---|---|---|---|---|---|---|
| Bachmair 2022[8] | HCPs in RA departments by telephones; supervised by exercise therapists | D | 5* | 45 | Physiotherapist | £267.15 |
| Turner 2016[9] | Tele-counselling conducted by the study therapists | D | 5<br>1 | 45†<br>90 | Physiotherapist | £294.89 |
| Katz 2018[10] | NR | D | 10 | NR | NE | NE |
| Kucharski 2019[11] | Physiotherapist | mixed | 1 | NR | Physiotherapist | NE |
| Torkhani 2021[12] | 15 min phone call by a specialised trainer in adapted physical activity | mixed | 8 | 15 | Physiotherapist | £112.34 |

*median 5 session was used.

† A session can last between 30 to 60 minutes.

HCP, health care professional; RA, rheumatoid arthritis; NR, not reported; NE, not estimable

**Table S 4** Costs of CBT-Fatigue delivered to individuals

| First author and year of publication | Session facilitator | Face to face (F) or distance (D) | Number of sessions | Duration of sessions (minutes) | Unit cost | Cost per patient |
|---|---|---|---|---|---|---|
| Mead 2022[13] | Non-psychology health care professionals (nurses and physiotherapists) | D | 7 | 60 | CBT | £902.68 |
| Ehde 2015[14] | Master-level social workers or doctoral-level psychologist | D | 8 | 60 | CBT | £1,096.12 |
| | | | 2 | 15 | CBT | |
| Artom 2019[15] | Therapist | D | 1 | 60 | CBT | £580.30 |
| | | | 7 | 30 | | |
| Nguyen 2019[16] | Three psychologists with doctoral qualifications in clinical neuropsychology and advanced training in CBT | F | 9 | NR | CBT | NE |
| Menting 2017[17] | Therapist | mixed | 5.4 | 50 | CBT | £580.30 |
| Moss-Morris 2012[18] | Internet-based intervention and assistant psychologist | D | 3 | 45* | CBT | £290.15 |
| Picariello 2021[19] | Psychologists | mixed | 2 | 60 | CBT | £451.34 |
| | | | 3 | 30 | | |
| Jhamb 2023[20] | CBT therapists | D | 12 | 52.5† | CBT | £1,354.03 |
| van den Akker 2017[21] | Psychologist (CBT Therapist) | F | 12.00 | 45 | CBT | £1,160.59 |
| van Kessel 2008[22] | Clinical psychologist | mixed | 8.00 | 50 | CBT | £859.70 |
| Okkersen 2018[23] | Therapists | mixed | 12.00 | NR | CBT | NE |
| Pottgen 2018[24] | Internet-based intervention | D | 8.00 | 60 | Unclear | NE |
| Bachmair 2022[8] | HCPs in RA departments by telephones; supervised by CBT/clinical psychologists | D | 8 | 45 | CBT | £773.73 |

*A telephone session can last between 30 to 60 minutes.

†A session can last between 45 to 60 minutes.

CBT, cognitive behavioural therapy; HCP, health care professional; RA, rheumatoid arthritis; NR, not reported; NE, not estimable

**Table S 5** Costs of mindfulness delivered to individuals

| First author and year of publication | Session facilitator | Face to face (F) or distance (D) | Number of sessions | Duration of sessions (minutes) | Unit cost | Cost per patient |
|---|---|---|---|---|---|---|
| Goren 2022[25] | Clinical social workers with a special training in CBT and mindfulness-based stress reduction | D | 7 | 60 | Mindfulness-based cognitive therapy | £724.03 |
| Torkhani 2021[12] | 15 min phone call by a specialised trainer in adapted physical activity | D | 8 | 15 | Mindfulness-based cognitive therapy | £206.87 |

CBT, cognitive behavioural therapy

**Table S 6** Costs of physical activity promotion delivered to groups

| First author and year of publication | Session facilitator | Face to face (F) or distance (D) | Number of sessions | Duration of sessions (minutes) | Total individuals started | Number of groups | Unit cost | Cost per patient |
|---|---|---|---|---|---|---|---|---|
| Lutz 2017[26] | Sport scientist (2 staff members per group) | F | 12 | 75* | 8 | 1 | Physiotherapist | £210.64 |
| Callahan 2014[27] | 17 instructors for 17 centres (assumed 1 instructor per group) | F | 20 | 60 | 172 | 17 | Physiotherapist | £102.13 |

*A session can last between 60 to 90 minutes.

**Table S 7** Costs of CBT-Fatigue delivered to groups

| First author and year of publication | Session facilitator | Face to face (F) or distance (D) | Number of sessions | Duration of sessions (minutes) | Total individuals started | Number of groups | Average number of persons per group | Unit cost | Cost per patient |
|---|---|---|---|---|---|---|---|---|---|
| Zedlitz 2012[28] | Psychologists for CBT | F | 12 | 120 | 45 | 12 | 4 | CBT | £773.73 |
| Gay 2023[29] | FACETS+ programme delivered by 2 psychologists | F | 10 | 90 | - | 1 | 8* | CBT | £483.58 |
| Hewlett 2019[30] | Rheumatology nurses and occupational therapists | F | 6 | 120 | - | - | 6 | CBT | £558.80 |
| | | | 1 | 60 | | | | | |
| Bredero 2023[31] | Mindfulness trainer | F | 8 | 150 | 47 | 3 | 16 | CBT | £378.63 |
| | | | 1 | 180 | | | | | |
| Thomas 2013[32] | Two health professionals (occupational therapists/MS specialist nurses/physio-therapists) | F | 6 | 90 | - | - | 9† | CBT | £257.91 |

*6 to 10 persons per group
†6 to 12 persons per group
CBT, cognitive behavioural therapy; MS, multiple sclerosis

**Table S 8** Costs of mindfulness delivered to groups

| First author and year of publication | Session facilitator | Face to face (F) or distance (D) | Number of sessions | Duration of sessions (minutes) | Total individuals started | Number of groups | Average number of persons per group | Unit cost | Cost per patient |
|---|---|---|---|---|---|---|---|---|---|
| Grossman 2010[33] | 2 experienced certified teachers with > 9 years of teaching experience. | F | 8 | 150 | - | - | 13* | Mindfulness-based cognitive therapy | £214.82 |
| | | | 1 | 420 | - | - | | | |

*10 to 15 persons per group

**Calculation of baseline fatigue score and standard deviation (FSS) for scenario analysis**

The network meta-analysis (NMA) presented elsewhere on the effect of non-pharmacological interventions for fatigue management used standardised mean difference of change of fatigue from baseline in order to manage the use of different fatigue scales. Studies which measured fatigue outcomes using the FSS score within the base case end of treatment network were used to evaluate the baseline FSS score. Baseline scores were pooled across all arms under the assumption that baseline fatigue should not significantly differ between study arms conditional on proper randomisation.

Forty study arms had baseline fatigue measured using FSS. Some studies were found to report a non-averaged FSS, and therefore were not included within the evaluation of the representative baseline FSS. The average baseline FSS score and the corresponding groups were evaluated using the formulae in Section 6.5.2.10 in the Cochrane Handbook for combining summary statistics across groups.[34]

The pooled baseline FSS score was 5.35 with a standard deviation (SD) of 1.16. As this was the average score across 9 domains, and the economic model uses total FSS score with its SD, these were scaled up by a factor of 9 for inclusion in the economic model.

**NMA scenario analysis, LT data follow-up time**

For studies that reported fatigue scores for two different long-term follow-ups, the base-case NMA included the longest follow-up data but a sensitivity analysis was conducted incorporating the earliest long-term follow-up point (i.e. closest to 3 months after EOT). Only 5 studies out of 18 studies including within the long-term NMA had more than one follow-up greater than 3 months. For studies without alternative data, the original data (as in the base case) was used.

The detailed methodology of the NMA, and included studies is included elsewhere.[35] For comparison, the predicted treatment effect point estimates for the interventions considered within the economic modelling, and their corresponding 95% credible intervals for the base case and the scenario analysis are compared in Table S9. Note that CBT-Fatigue is considered as a combination of both group and individual interventions.

**Table S 9** Comparison of base case predicted treatment effects for three interventions within the economic model with the predicted treatment effects when using long-term follow-up data closest to 3 months, as opposed to maximum available follow-up

| **Predicted treatment effect from NMA (LT)** | | |
|---|---|---|
| Intervention | Base case point, estimate (95% CrI) [Number of studies] | Use of maximum follow-up within LT, point estimate (95% CrI) [Number of studies] |
| Mindfulness based | -0.54 (-0.99, -0.105) [1] | -0.54 (-1.056, -0.035) [1] |
| Physical activity promotion | -0.52 (-0.862, -0.184) [2] | -0.53 (-0.928, -0.125) [2] |
| CBT-Fatigue | -0.4 (-0.627, -0.214) [9] | -0.42 (-0.684, -0.196) [9] |

CrI, credible interval; NMA, network meta-analysis; LT, long-term; CBT-Fatigue, cognitive behavioural therapy for fatigue

Generally there was minimal difference in the point estimates of the predicted treatment effects. However, there was some broadening of the 95% credible intervals (CrIs) , likely due to the broader range of follow-up times included within the scenario analysis. Despite this, the three interventions remained statistically significant.

**Comparison of clinical effectiveness for CBT for fatigue in group versus individual delivery setting**

Within the clinical effectiveness comparisons, published elsewhere, data was recorded whether interventions were administered in a group or individual setting. For CBT-Fatigue, multiple studies at ST and LT were recorded to have used group and individual based interventions. Therefore, the comparative efficacy of CBT-Fatigue (group) and CBT-Fatigue (individual) could be assessed in a scenario analysis at EOT, ST, and LT follow-up.

The detailed methodology of the network meta-analyses is published elsewhere. Here, the predicted treatment effect point estimate and associated 95% CrIs are presented for CBT-Fatigue (group), CBT-Fatigue (individual) and pooled CBT-Fatigue (from the base case) for each follow-up analysis. Other intervention treatment effects are not included, but there was minimal impact on the predicted treatments effects for other interventions.

**Table S 10** Comparison of CBT-Fatigue (group), CBT-Fatigue (individual) and CBT-Fatigue (pooled) predicted treatment effects at EOT, ST and LT

| **Predicted treatment effect for CBT-Fatigue (group) vs (individual)** | | | |
|---|---|---|---|
| Intervention | EOT<br>Point estimate (95% CrI)<br>[Number of studies] | ST<br>Point estimate (95% CrI)<br>[Number of studies] | LT<br>Point estimate (95% CrI)<br>[Number of studies] |
| CBT-Fatigue (group) | -0.60 (-1.00, -0.192) [4] | -0.06 (-0.486, 0.371) [1] | -0.38 (-0.679, -0.128) [4] |
| CBT-Fatigue (individual) | -0.64 (-0.898, -0.395) [13] | -0.23 (-0.57, 0.071) [6] | -0.47 (-0.93, -0.046) [5] |
| CBT-Fatigue (pooled) | -0.63 (-0.865, -0.402) [17] | -0.17 (-0.415, 0.062) [7] | -0.4 (-0.627, -0.214) [9] |

CrI, credible interval; NMA, network meta-analysis; EOT, end of treatment; ST, short-term; LT, long-term; CBT-Fatigue, cognitive behavioural therapy for fatigue

Noticeably, there were fewer studies influencing CBT-Fatigue nodes within the NMA at ST compared to EOT and LT, corresponding predicted treatment effects were all reduced and found not to be statistically significant. At both EOT and ST there were more studies implementing individual CBT-Fatigue, as opposed to group-based interventions. The 95% credible intervals of group and individual interventions overlapped at all follow-up times, suggesting no meaningful differences in predicted treatment effects. However, there were moderate differences in the ST estimates, with CBT-Fatigue in a group setting being shown to be potentially less beneficial than in the individual setting, however this was evidenced by a single study only, and this was not subsequently evident at LT follow-up.

**Table S 11** Input parameters for the base case analysis

| Parameter | Value | Standard error (SE) | Distribution | Source |
|---|---|---|---|---|
| **SMD of interventions (clinical effectiveness), EOT network** | | | | |
| Usual care | 0.00 | - | Fixed | Systematic review and NMA[35] |
| Physical activity promotion | -0.32 | - | CODA | |
| CBT-Fatigue-I | -0.63 | - | CODA | |
| CBT-Fatigue-G | -0.63 | - | CODA | |
| Mindfulness | -0.59 | - | CODA | |
| **SMD of interventions (clinical effectiveness), ST network** | | | | |
| Usual care | 0.00 | - | Fixed | Systematic review and NMA[35] |
| Physical activity promotion | -0.51 | - | CODA | |
| CBT-Fatigue-I | -0.17 | - | CODA | |
| CBT-Fatigue-G | -0.17 | - | CODA | |
| Mindfulness | - | - | | |
| **SMD of interventions (clinical effectiveness), LT network** | | | | |
| Usual care | 0.00 | - | Fixed | Systematic review and NMA[35] |
| Physical activity promotion | -0.52 | - | CODA | |
| CBT-Fatigue-I | -0.40 | - | CODA | |
| CBT-Fatigue-G | -0.40 | - | CODA | |
| Mindfulness | -0.54 | - | CODA | |
| **Expected costs per patient (individual interventions)** | | | | |
| Usual care | £0 | - | Fixed | Assumption |
| Physical activity promotion | £267 | 46.57 | Gamma | Systematic review and NMA[35] |
| CBT-Fatigue-I | £817 | 271.40 | Gamma | |
| Mindfulness | £465 | 131.93 | Gamma | |
| **Expected costs per patient (group intervention)** | | | | |
| CBT-Fatigue-G | £484 | 131.587 | Gamma | Systematic review and NMA[35] |
| Physical activity promotion | £156 | 27.681 | Gamma | |
| Mindfulness | £215 | 53.705 | Gamma | |
| **Mapping parameters** | | | | |
| Baseline FSS scores | 43.73 | 15.10 | Normal | Goodwin *et al.*2019[3] |
| Mapping coefficient_constant | 0.8970 | 0.015 | Normal | |
| Mapping coefficient_ FSS | 0.006000 | 0.0003 | Normal | |
| **Average timepoints (months)** | | | | |
| **Baseline to EOT** | | | | |
| Usual care | 3.26 | - | Fixed | NMA[35] |
| Physical activity promotion | 4.04 | - | Fixed | |
| CBT-Fatigue-I | 3.07 | - | Fixed | |
| CBT-Fatigue-G | 3.07 | - | Fixed | |
| Mindfulness | 2.23 | - | Fixed | |

| | | | | |
|---|---|---|---|---|
| **EOT to ST** | | | | |
| Usual care | 2.23 | - | Fixed | NMA[35] |
| Physical activity promotion | 1.38 | - | Fixed | |
| CBT-Fatigue-I | 1.91 | - | Fixed | |
| CBT-Fatigue-G | 1.91 | - | Fixed | |
| Mindfulness | 0.00 | - | Fixed | |
| **ST to LT** | | | | |
| Usual care | 5.85 | - | Fixed | NMA[35] |
| Physical activity promotion | 8.53 | - | Fixed | |
| CBT-Fatigue-I | 6.45 | - | Fixed | |
| CBT-Fatigue-G | 6.45 | - | Fixed | |
| Mindfulness | 6.00 | - | Fixed | |
| **LT to final end points** | | | | |
| Usual care | 12.66 | - | Fixed | NMA[35] |
| Physical activity promotion | 10.05 | - | Fixed | |
| CBT-Fatigue-I | 12.58 | - | Fixed | |
| CBT-Fatigue-G | 12.58 | - | Fixed | |
| Mindfulness | 15.77 | - | Fixed | |

SMD, standardised mean difference; EOT, end of treatment; ST, short-term; LT, long-term; CBT-Fatigue-I, cognitive behavioural therapy for fatigue (individual); CBT-Fatigue-G, cognitive behavioural therapy for fatigue (group); FSS, fatigue severity score; CODA, Convergence Diagnostic and Output Analysis; NMA, network meta-analysis

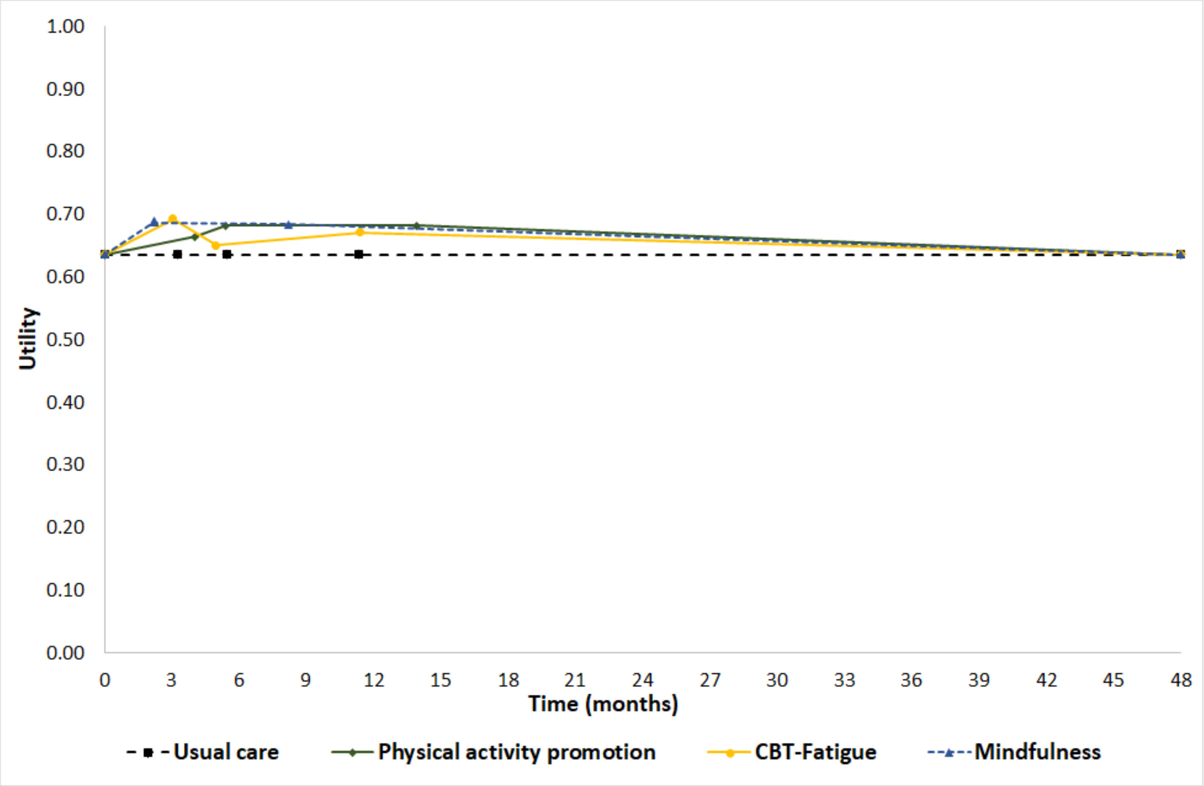


**Figure S 1** Utility values of interventions across different timepoints (optimistic scenario)

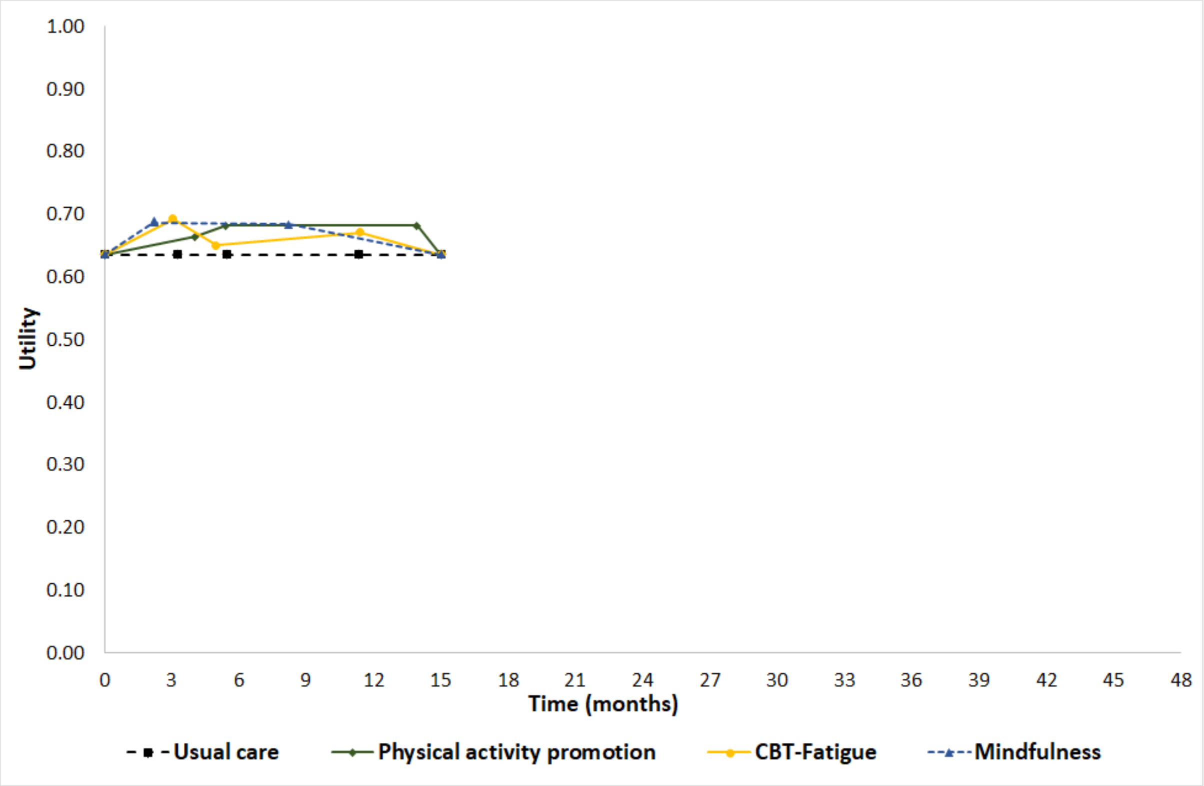


**Figure S 2** Utility values of interventions across different timepoints (pessimistic scenario)

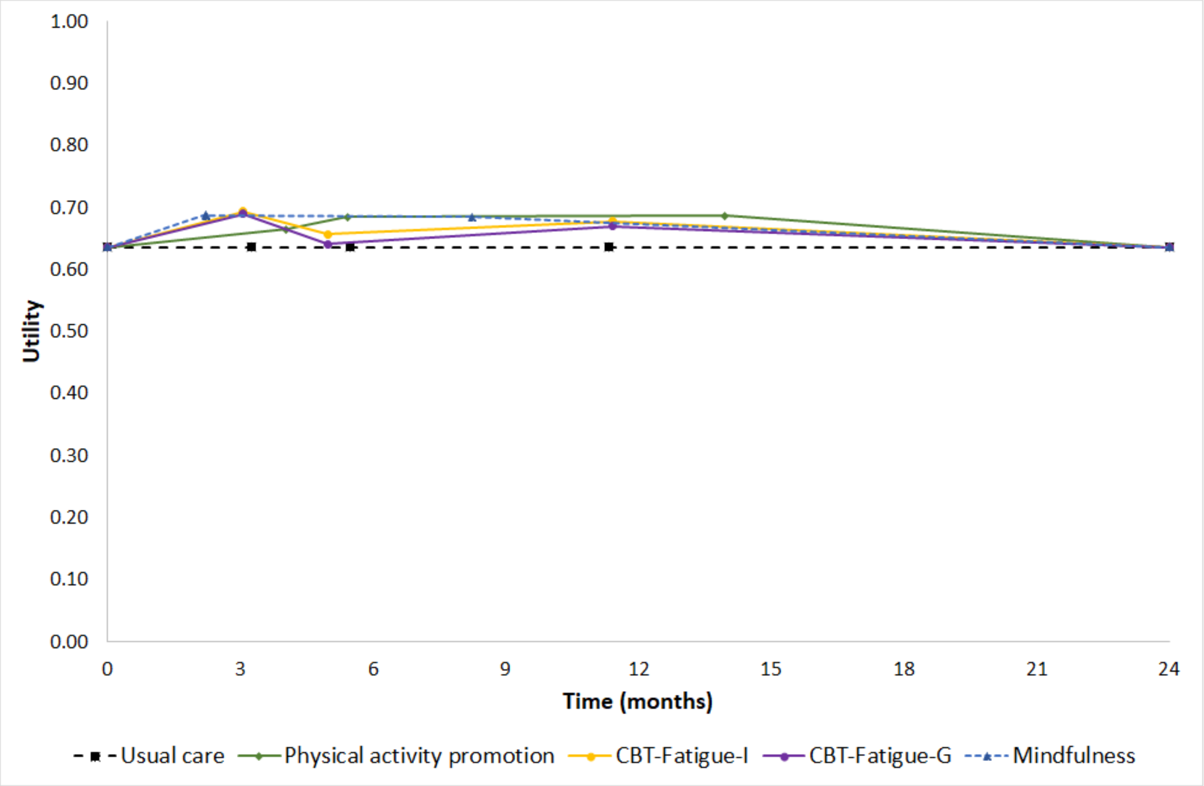


**Figure S 3** Utility values of interventions across different timepoints (using results from NMA scenario analysis 1 comparing individual vs group CBT-Fatigue)

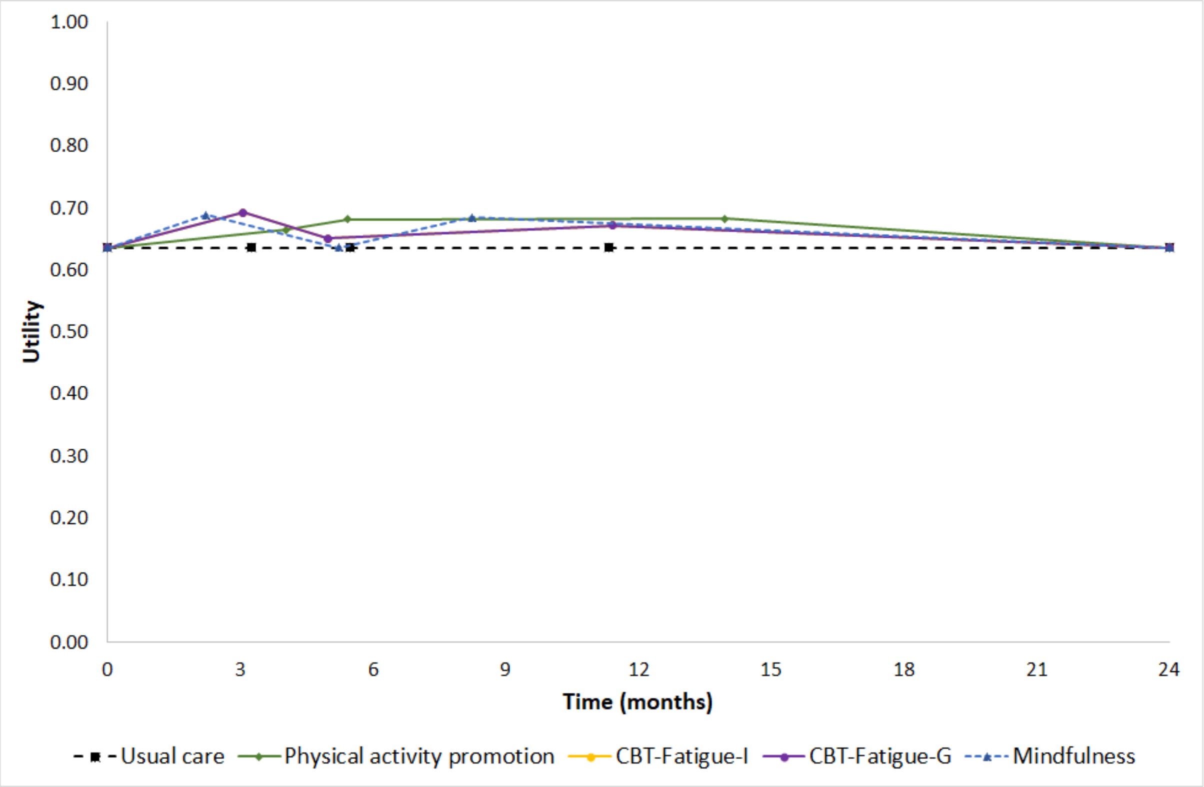


**Figure S 4** Utility values of interventions across different timepoints (removal of the linear change assumption for mindfulness at the short-term follow-up timepoint)

**Table S 12** Results of scenario analyses (probabilistic)

| Intervention type | Group or individual intervention | Median cost (£) | Mean QALYs (discounted) | Incremental Costs | Incremental QALYs | NMB at £20,000 threshold | INMB against UC |
|---|---|---|---|---|---|---|---|
| **Using optimistic assumption for the duration of treatment effect decline (48 months from the baseline)** | | | | | | | |
| Usual care | - | £0 | 2.364 | - | - | £47,286 | - |
| Physical activity promotion | Individual | £267 | 2.467 | £267 | 0.103 | £49,079 | £1,793 |
| CBT-Fatigue | Individual | £810 | 2.443 | £810 | 0.078 | £48,043 | £758 |
| Mindfulness | Individual | £462 | 2.470 | £462 | 0.106 | £48,943 | £1,658 |
| Physical activity promotion | Group | £157 | 2.467 | £157 | 0.103 | £49,190 | £1,904 |
| CBT-Fatigue | Group | £485 | 2.443 | £485 | 0.078 | £48,369 | £1,083 |
| Mindfulness | Group | £214 | 2.470 | £214 | 0.106 | £49,191 | £1,905 |
| **Using pessimistic assumption for the duration of treatment effect decline (15 months from the baseline)** | | | | | | | |
| Usual care | - | £0 | 0.774 | - | - | £15,482 | - |
| Physical activity promotion | Individual | £267 | 0.817 | £267 | 0.043 | £16,079 | £598 |
| CBT-Fatigue | Individual | £810 | 0.806 | £810 | 0.032 | £15,310 | -£172 |
| Mindfulness | Individual | £462 | 0.818 | £462 | 0.043 | £15,889 | £407 |
| Physical activity promotion | Group | £157 | 0.817 | £157 | 0.043 | £16,190 | £709 |
| CBT-Fatigue | Group | £485 | 0.806 | £485 | 0.032 | £15,636 | £154 |
| Mindfulness | Group | £214 | 0.818 | £214 | 0.043 | £16,137 | £655 |
| **Using baseline FSS score and SE from EOT NMA network** | | | | | | | |
| Usual care | - | £0 | 1.172 | - | - | £23,447 | - |
| Physical activity promotion | Individual | £267 | 1.214 | £267 | 0.042 | £24,013 | £566 |
| CBT-Fatigue | Individual | £810 | 1.204 | £810 | 0.031 | £23,261 | -£186 |
| Mindfulness | Individual | £462 | 1.215 | £462 | 0.042 | £23,832 | £385 |
| Physical activity promotion | Group | £157 | 1.214 | £157 | 0.042 | £24,124 | £677 |
| CBT-Fatigue | Group | £485 | 1.204 | £485 | 0.031 | £23,587 | £139 |
| Mindfulness | Group | £214 | 1.215 | £214 | 0.042 | £24,080 | £633 |
| **Using different SMDs between individual and group CBT-Fatigue interventions** | | | | | | | |
| Usual care | - | £0 | 1.223 | - | - | £24,467 | - |
| Physical activity promotion | Individual | £267 | 1.287 | £267 | 0.064 | £25,472 | £1,005 |
| CBT-Fatigue | Individual | £810 | 1.275 | £810 | 0.052 | £24,696 | £229 |
| Mindfulness | Individual | £462 | 1.284 | £462 | 0.060 | £25,212 | £745 |

| Intervention type | Group or individual intervention | Median cost (£) | Mean QALYs (discounted) | Incremental Costs | Incremental QALYs | NMB at £20,000 threshold | INMB against UC |
|---|---|---|---|---|---|---|---|
| Physical activity promotion | Group | £157 | 1.287 | £157 | 0.064 | £25,583 | £1,116 |
| CBT-Fatigue | Group | £485 | 1.263 | £485 | 0.040 | £24,773 | £306 |
| Mindfulness | Group | £214 | 1.284 | £214 | 0.060 | £25,460 | £993 |
| **Using alternative SMDs using the shorter follow-up point in the LT NMA network** | | | | | | | |
| Usual care | - | £0 | 1.224 | - | - | £24,474 | - |
| Physical activity promotion | Individual | £267 | 1.284 | £267 | 0.060 | £25,406 | £932 |
| CBT-Fatigue | Individual | £810 | 1.270 | £810 | 0.046 | £24,584 | £110 |
| Mindfulness | Individual | £462 | 1.285 | £462 | 0.061 | £25,228 | £754 |
| Physical activity promotion | Group | £157 | 1.284 | £157 | 0.060 | £25,517 | £1,043 |
| CBT-Fatigue | Group | £485 | 1.270 | £485 | 0.046 | £24,909 | £435 |
| Mindfulness | Group | £214 | 1.285 | £214 | 0.061 | £25,476 | £1,002 |
| **Assuming the SMD of mindfulness at the ST follow-up timepoint is zero** | | | | | | | |
| Usual care | - | £0 | 1.224 | - | - | £24,471 | - |
| Physical activity promotion | Individual | £267 | 1.284 | £267 | 0.060 | £25,404 | £934 |
| CBT-Fatigue | Individual | £810 | 1.269 | £810 | 0.045 | £24,560 | £90 |
| Mindfulness | Individual | £462 | 1.272 | £462 | 0.049 | £24,985 | £514 |
| Physical activity promotion | Group | £157 | 1.284 | £157 | 0.060 | £25,515 | £1,044 |
| CBT-Fatigue | Group | £485 | 1.269 | £485 | 0.045 | £24,886 | £415 |
| Mindfulness | Group | £214 | 1.272 | £214 | 0.049 | £25,233 | £762 |
| **Using the lowest cost of intervention across studies** | | | | | | | |
| Usual care | - | £0 | 1.225 | - | - | £24,492 | - |
| Physical activity promotion | Individual | £113 | 1.285 | £113 | 0.060 | £25,578 | £1,085 |
| CBT-Fatigue | Individual | £285 | 1.270 | £285 | 0.045 | £25,106 | £613 |
| Mindfulness | Individual | £203 | 1.286 | £203 | 0.061 | £25,507 | £1,015 |
| Physical activity promotion | Group | £102 | 1.285 | £102 | 0.060 | £25,588 | £1,096 |
| CBT-Fatigue | Group | £259 | 1.270 | £259 | 0.045 | £25,131 | £639 |
| Mindfulness | Group | £161 | 1.286 | £161 | 0.061 | £25,550 | £1,058 |
| **Using the highest cost of intervention across studies** | | | | | | | |
| Usual care | - | £0 | 1.223 | - | - | £24,464 | - |
| Physical activity promotion | Individual | £295 | 1.283 | £295 | 0.060 | £25,371 | £906 |
| CBT-Fatigue | Individual | £1,348 | 1.268 | £1,348 | 0.045 | £24,017 | -£447 |

| Intervention type | Group or individual intervention | Median cost (£) | Mean QALYs (discounted) | Incremental Costs | Incremental QALYs | NMB at £20,000 threshold | INMB against UC |
|---|---|---|---|---|---|---|---|
| Mindfulness | Individual | £721 | 1.284 | £721 | 0.061 | £24,965 | £501 |
| Physical activity promotion | Group | £211 | 1.283 | £211 | 0.060 | £25,455 | £991 |
| CBT-Fatigue | Group | £775 | 1.268 | £775 | 0.045 | £24,590 | £125 |
| Mindfulness | Group | £268 | 1.284 | £268 | 0.061 | £25,418 | £953 |

QALY, quality-adjusted life years; NMB, net monetary benefit; INMB, incremental net monetary benefit; UC, usual care; CBT, cognitive behavioural therapy; EOT: end-of-treatment; LT, long-term; NMA, network meta-analysis; FSS, fatigue severity score; SE, standard error; SMD, standardised mean difference

**Original protocol of the study**

The original protocol of the EIFFEL study is reported here:
https://www.fundingawards.nihr.ac.uk/award/NIHR154660
